# Blood DNA Methylation Predicts Long-Term Risk of Dementia in Prospective Cohorts

**DOI:** 10.64898/2026.07.08.26357554

**Authors:** Adam X. Maihofer, Caroline E. Mackey, Josephine A. Robertson, Riccardo E. Marioni, Steve Nguyen, Linda K. McEvoy, Andrea Z. LaCroix, Mark A. Espeland, Stephen R. Rapp, Susan M. Resnick, Bowei Zhang, Steve Horvath, Kenneth Beckman, Towia A. Libermann, Tom C. Russ, Simon R. Cox, Sarah E. Harris, Maira Pyrgioti, Aladdin H. Shadyab

## Abstract

Blood-based DNA methylation may help identify biological changes related to dementia risk before clinical symptoms appear. We conducted an epigenome-wide association study of incident all-cause dementia in 5,999 cognitively healthy women from the Women’s Health Initiative Memory Study, 777 of whom developed dementia over up to 25 years of follow-up. Baseline blood DNA methylation was tested for association with time to dementia. One CpG site, cg05917797, was significantly associated with dementia risk, with higher methylation linked to lower risk. This association was only minimally changed after accounting for APOE ε4 carrier status, plasma p-tau217, and epigenetic aging measures. cg05917797 also replicated in meta-analysis of four independent prospective cohorts including 10,916 participants and 413 incident dementia cases. In post-mortem brain methylation datasets, higher methylation at cg05917797 was associated with lower Braak stage in temporal gyrus and cerebellum. In meta-analysis of all five prospective cohorts, including 16,915 participants and 1,190 dementia cases, cg05917797 remained the leading association, and three additional CpGs were identified for further study. These findings support cg05917797 as a reproducible blood-based epigenetic marker of long-term dementia risk.

## INTRODUCTION

Worldwide, 46.8 million people have dementia, which is expected to increase to 131.5 million by 2050.^1^ While the genetic basis of dementia has been extensively studied, there is increasing recognition that epigenetic mechanisms may also play a role in dementia pathophysiology.^2^ And indeed, unlike static genetic variants, which have been extensively studied, DNA methylation (DNAm) is responsive to environmental, lifestyle, and pathological factors, making it a promising candidate for identifying biomarkers that reflect early and potentially modifiable processes in dementia pathogenesis.^3^

To date, a substantial number of blood and brain DNA methylation (DNAm) studies of dementia have been conducted. However, many have used cross-sectional or case-control designs with small sample sizes, precluding understanding of the DNAm differences that precede onset of dementia among cognitively healthy individuals.^4–13^ Thus, an unresolved question is whether peripheral DNA methylation captures neurodegenerative risk states long before clinical onset, rather than reflecting consequences of disease progression closer to diagnosis. More recent, longitudinal studies of incident dementia have been conducted to help address this question.^14,15^ They have identified several CpG sites associated with time to dementia, as well as implicated specific biological pathways, including metabolic and immune pathways. However, results are not consistent across studies, which is potentially attributable to factors such as limitations of discoverability at given sample sizes or heterogeneity across cohorts.

To address these limitations, we performed the largest epigenome-wide association study (EWAS) of incident all-cause dementia to date. We examined 5,999 participants from the Women’s Health Initiative Memory Study (WHIMS) who were cognitively healthy at baseline, with 777 cases of incident dementia during an average 11-year follow-up. We performed replication and an extensive multi-omic analysis to elucidate the mechanisms underlying the observed associations. To enhance discovery power to identify significant DNAm-dementia associations, we performed a meta-analytic EWAS of incident dementia among five prospective cohorts including WHIMS, Framingham Heart Study Exam 9, Alzheimer’s Disease Neuroimaging Initiative, Generation Scotland, and Lothian Birth Cohort 1936, amounting to a total sample size of 16,915 participants with 1,190 cases of incident dementia.

Here, we show that blood DNA methylation measured in cognitively healthy older adults captures long-term risk of dementia, identifying a single CpG site that predicts incident dementia up to two decades before diagnosis and replicates across independent cohorts and brain-based datasets. Our meta-analysis of all cohorts identified additional CpG sites for future follow-up investigations.

## RESULTS

### Baseline Characteristics

Participants were prospectively assessed for dementia.^16,17^ DNA methylation was measured in blood at baseline using the Illumina EPIC V2 array. Following quality control procedures, the analytic sample from WHIMS included 5,999 women who were dementia-free at baseline (Table 1). The average age at baseline was 70 years old. The majority of participants (86%) were of European Ancestry, with the next largest group being of African Ancestry (6%). During follow-up, 13% of participants (N=777) developed dementia. Compared to women who did not develop dementia, those with incident dementia were slightly older at baseline (0.8 years, p= 8.4x10^-9^) and had longer follow-up (14 years versus 11.1 years, p=4.8x10^-34^).

**Table 1.**
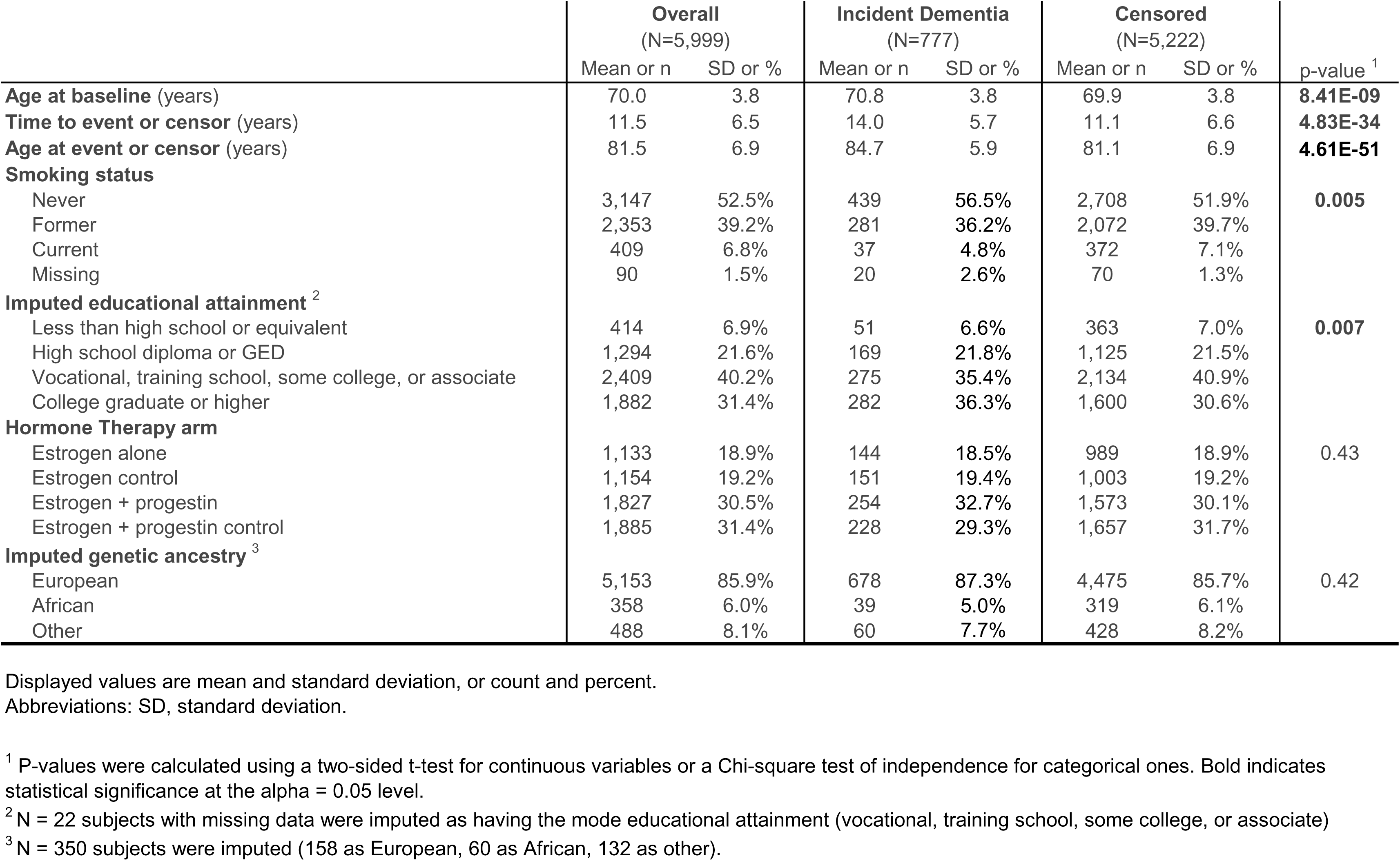
Demographics of WHIMs participants.

There were significant differences in educational attainment (p=0.007): cases were more likely to have graduated college or higher education, and controls were more likely to have had vocational training, training school, some college, or an associate’s degree, although differences were modest (we note this association attenuates to non-significance upon adjustment for other covariates in multivariable models). Further, there was a significant difference in smoking history (p=0.005), where relative to controls, cases were more likely to have never smoked or be former smokers; however, differences were modest. There were no significant differences in hormone therapy (HT) arm (p=0.43). There were no significant differences in genetic ancestry (European, African, Latin American, Asian, or ‘Other’ ancestry groups) (p=0.42), but there was a significant difference in the more finely grained ancestry PC2 (p=0.01). There were no significant differences in cell type proportion or chip-row position (Supplementary Table 1).

### Epigenome-wide association study (EWAS) of time to dementia

We assessed the association between baseline DNA methylation at N=870,459 CpG sites and incident dementia (Figure 1; EWAS summary data in Supplementary Data 1). There was minimal inflation of test statistics (GC lambda = 1.02; Supplementary Figure 1), suggesting limited influence of residual technical artifacts on results.

**Figure 1.**
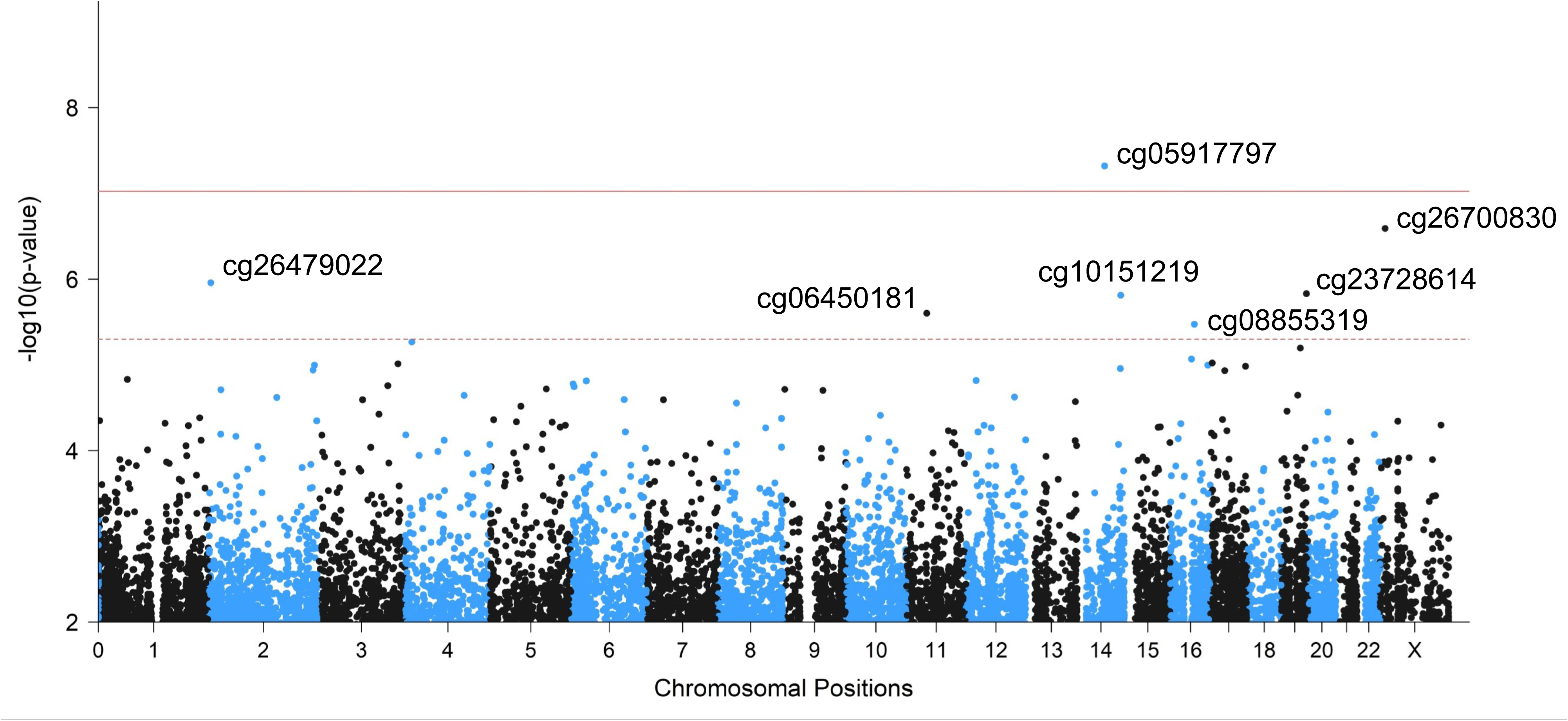
Manhattan plot of incident dementia EWAS in WHIMS. The solid red line indicates genome-wide significance (p < 9x10^-8^) and the dotted red line indicates suggestive association (p < 5x10^-6^)

A single CpG site was epigenome-wide significantly associated with incident dementia (cg05917797; chr14:62,075,110 BP; logHR = -0.7744, SE =0.1419, p=4.8x10^-8^; Supplementary Figure 2 for data distribution). As dementia incidence was on average 11 years (and up to 25 years) from baseline, it was consistent with cg05917797 having a potential role as an early marker of disease susceptibility rather than a correlate of symptomatic disease (Figure 2 survival analysis).

**Figure 2.**
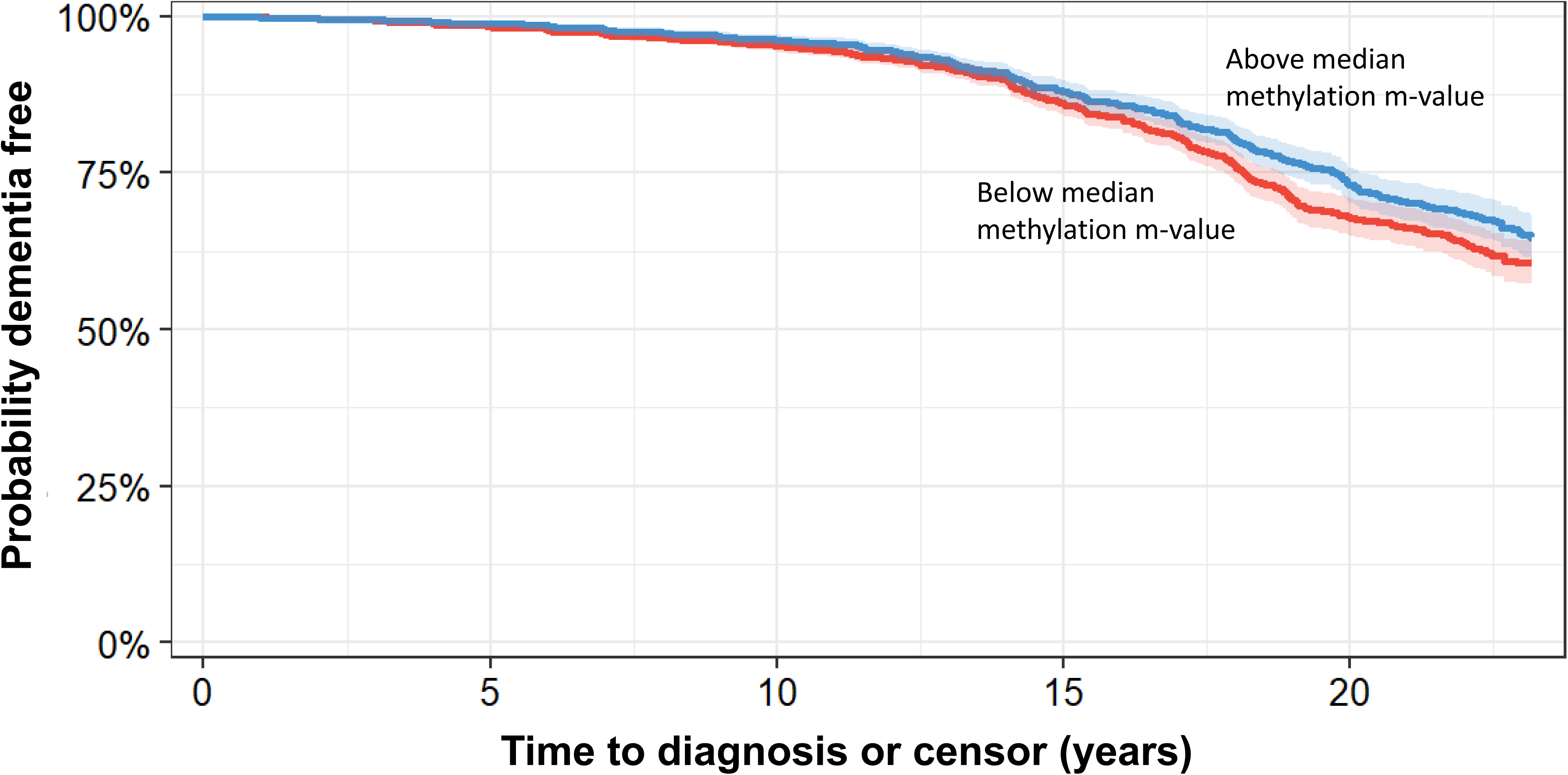
Kaplan-Meier curves of dementia-free survival by cg05917797 methylation m-values (above vs. below median). The median cg05917797 methylation m-value was4.66. Shading represents confidence intervals. Capped follow-up at the 95th percentile (23.2 years) due to smaller risk sets from censoring and unstable survival estimates. (N=5,996 WHIMS subjects)

Six other CpGs were suggestively associated with incident dementia (defined as p < 5x10^-6^; Supplementary Table 2 of suggestive loci across our and other dementia cohorts). We next sought to contextualize general association signals identified in our EWAS through pathway analysis in GOmeth. Analysis of the top 1000 most significant CpG sites identified one significant pathway, “Integrin Signaling” (containing N=153 CpGs, with 18 being differentially methylated, pathway p = 1.17x10^-4^ and FDR=0.043).

For sensitivity analyses of the overall EWAS, we repeated the analyses with: 1) additional demographic covariates that were significantly different or may otherwise confound associations (education, HT randomization arm, and smoking history) to augment the base set of covariates (Supplementary Data 2); and 2) restriction to only European ancestry samples (Supplementary Data 3). In both cases, the overall results were substantively the same; the overall effect size correlations with the primary result were 99% and 99%, respectively. In particular, there was little change (<10%) in the reported effect size of cg05917797 for the models that included these additional covariates (Supplementary Table 3). The Cox proportional hazards assumption for cg05917797 was satisfied (global proportional hazards test p=0.90). We performed a Cox model that accounted for death as a competing risk, for which there was little difference in the effect size of cg05917797 (cause specific Cox model log[HR]=-0.7217, SE=0.1389, p=2.0x10^-7^) relative to the standard model.

### Independence of cg05917797 from known risk biomarkers of dementia

We attempted to delineate whether cg05917797 was capturing the effects of other known risk biomarkers, i.e. *APOE* e4 carrier status, epigenetic clock measures of accelerated aging, and plasma p-tau217 levels. We used each of these to predict cg05917797 methylation in models that also adjusted for our standard set of covariates. We observed nominal associations between cg05917797 and p-tau217 (N=2,371 samples with p-tau217 measured; Beta = -0.0128, SE = 0.0064, t = -1.952, p = 0.051), as well as between cg05917797 and *APOE* e4 carrier status (N = 5,112 with carrier status imputed, Beta = -2.071e-02, SE = 8.189e-03 t = -2.530 p = 0.011). In particular, we observed that these associations may be more specific to European ancestry participants, as the p-tau217 association was stronger in this subset (N=1,648; Beta =-2.493e-02, SE = 7.784e-03, t = -3.203, p = 0.001) and APOE carrier status was only reliably imputed in the European ancestry subset. We did not observe any association between the CpG and either epigenetic clock measure (PCGrimAge and AgeAccelGrim2; all p > 0.05).

Next, we evaluated whether adjustment for any of these biomarkers would result in attenuation of the association of cg05917797 with dementia risk. The association between cg05917797 and dementia was not substantially attenuated by adjustment for these factors (<15% change in estimate; Supplementary Table 4). These analyses suggest that cg05917797 captures a risk axis that is at least partially independent of established genetic and biomarker pathways.

### Replication of cg05917797 finding in external cohorts

For independent replication, we meta-analyzed four independent EWAS of incident dementia including the Alzheimer’s Disease Neuroimaging Initiative, Framingham Heart Study Exam 9, Generation Scotland, and Lothian Birth Cohort 1936 (N=10,916, including 413 incident dementia events). Our leading CpG site cg05917797 was replicated in the meta-analysis (log[HR] = -0.1881, SE = 0.0671, p=0.005; Figure 3). The consistency of this association across cohorts with differing designs, demographic structures, and follow-up durations supports the robustness of cg05917797 as a cross-cohort epigenetic signal of dementia risk.

**Figure 3.**
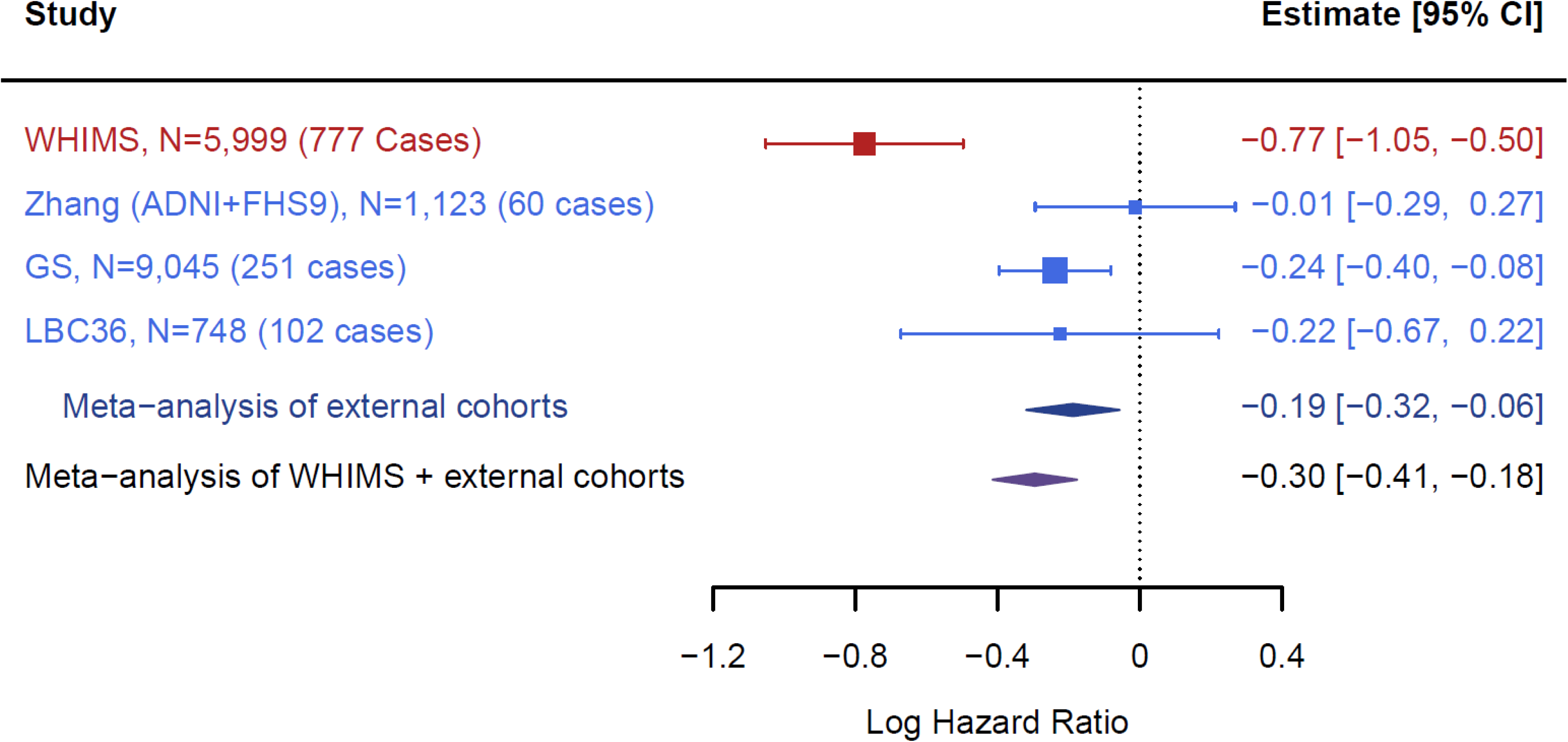
Meta-analysis of dementia EWAS at cg05917797. External cohorts (blue text, polygons) were meta-analyzed. Following this, they were meta-analyzed along with the WHIMS cohort (red text, polygons).

### Replication of cg05917797 in external brain tissues datasets

The relevance of brain tissues to dementia led us to question whether this association would also be present in external brain-based EWAS of dementia or a closely related pathology. A meta-analysis of post-mortem Alzheimer’s disease cohorts (N=1,453) examined the association of DNA methylation with Braak staging in three cortical regions and the cerebellum.^6^ The analysis identified cg05917797 as being associated with Braak stage in the superior or middle temporal gyrus (IVW Beta = -.0102, SE = 0.002, z = -5.19, p = 2.1x10^-7^) and cerebellum (IVW Beta = -0.00545, SE= 0.00124, Z = -4.38, p = 1.18x10^-5^) (Figure 4). Meta-analysis did not indicate significant associations in the prefrontal cortex (IVW Beta=0.0018, SE = 0.0017, p=0.294) or in the entorhinal cortex (IVW Beta = -0.0006, SE = 0.0039, p = 0.873).

**Figure 4.**
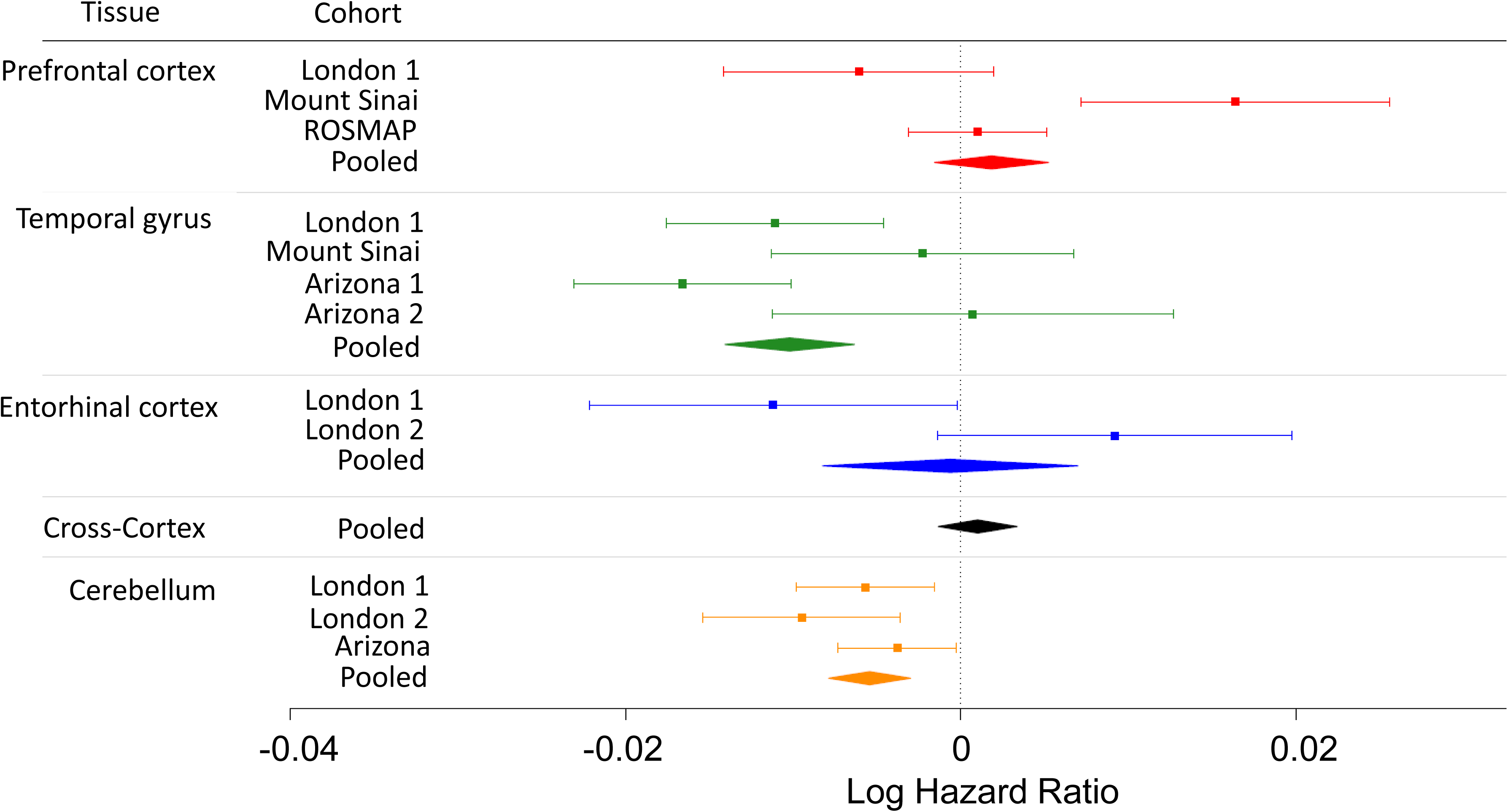
Meta analysis of Braak scores in different tissues and across cohorts for cg05917797. Results were inverse variance weighted meta-analyzed, as stratified by tissue type.

While these observations suggest relevance to brain biology, brain methylation measurements were not available in WHIMS, preventing direct evaluation of whether cg05917797 acts as a surrogate marker for brain methylation. We therefore examined external blood–brain methylation correlation resources, including the Blood Brain DNA Methylation Comparison Tool and BECon. Across datasets, cg05917797 was consistently hypermethylated in blood (mean methylation generally >0.9), as we had also observed within WHIMS. In contrast, methylation levels in brain tissue were substantially lower and more variable, with intermediate methylation observed across cortical regions including the prefrontal cortex, entorhinal cortex, superior temporal gyrus, and Brodmann areas 7, 10, and 20, and lower methylation observed in the cerebellum. Reported blood–brain correlations at cg05917797 were modest (absolute value of r < 0.3; Supplementary Figure 3 and 4). However, interpretation of these estimates is limited by the narrow range of blood methylation values observed at this CpG across reference datasets, which restricts the ability to reliably estimate cross-tissue correlation. Consequently, currently available reference datasets do not allow determination of whether disease-associated blood methylation differences at cg05917797 correspond to methylation differences in brain tissue, particularly in the context of dementia cases where we observed lower-than-average blood methylation at this otherwise highly methylated locus.

### Association of cg05917797 with cognitive function

We evaluated associations of cg05917797 with cognitive function in a subset of 1,910 WHIMS participants who had undergone longitudinal cognitive assessments (Supplementary Table 5). Higher levels of cg05917797 were associated with better average performance on short-delay free recall (Beta = 0.61, SE = 0.24, df=1887, t=2.60, p=0.0095), long delay free recall (Beta=0.62, SE=0.24, df=1887, t=2.62, p=0.0088), and forward digit span tasks (Beta=0.35, SE=0.16, df=1887, t=2.17, p=0.03). No significant associations were observed between methylation and PMAVoc total score, finger tapping in the dominant and nondominant hands, backward digit span, category fluency, or letter fluency (Supplementary Table 5). The observed associations attenuated to non-significance after adjustment for educational attainment (all p > 0.05). We also assessed whether cg05917797 was associated with cognitive decline using time interaction models but did not identify any significant time interaction (all p > 0.05).

Given associations with incident dementia and baseline cognitive performance, we explored whether cg05917797 was associated with cognitive traits in Generation Scotland, exploring the associations in the Generation Scotland cohort EWAS of cognitive abilities (N=9,162) (PMID: 35039062). They report directionally consistent, but relatively modest (all posterior importance probabilities < 0.05), associations of cg05917797 with cognitive function across various cognitive abilities measured: general cognitive ability (Beta = 1.49x10-4, SE=4.6x10^-5^, z=3.25, p=0.0012); vocabulary (Beta=1.31x10^-4^, SE = 4.68x10^-5^, z=2.86, p = 0.0042); and verbal fluency (Beta = 4.89x10^-5^, SE=2.46x10^-5^, z=1.199, p=0.0466). Other variable associations examined, while directionally concordant, were not significant, including general fluid cognitive ability (Beta = 2.14x10^-5^,SE=1.9x10-5,z=1.11,p=0.24), logical memory (Beta = 1.42x10^-5^, SE=1.2x10^-5^, z=1.18, p=0.27), and digit symbol score (Beta= 1.43x10^-5^, SE=1.6, z=0.89, p=0.37).

### Minimal impact of heritable genetic variation on cg05917797 methylation

mQTL studies suggest that methylation of some AD-related CpG sites may be influenced by genetic variation.^18,19^ We initially explored potential genetic influence on cg05917797 through lookup of methylation quantitative trait loci (mQTL) from GoDMC.^20^ While this did not display any significant associations, given multiple-comparison burden limits placed on published associations, it remained possible that meaningful associations exist. Accordingly, we also examined associations directly in WHIMS, performing a GWAS (N=3,860) in a subset of participants with quality controlled and imputed genotype data. This GWAS did not identify any genome-wide significant mQTLs, nor did any cis-SNP reach a suggestive level of association. Furthermore, SNP based heritability was not significantly different from zero, also suggesting minimal aggregate effect of SNPs with relatively weak-moderate effects.

### Plasma proteomic associations with cg05917797

To gain insights into biological pathways underlying cg05917797 methylation, we measured the association between cg05917797 and plasma abundance of 10,778 proteins in a subset of 2,390 WHIMS participants with SomaScan proteomics measured at the same time point. No protein was significant after adjustment for multiple comparisons (Supplementary Table 6), nor did pathway analysis of the leading associations (all proteins with p < 0.01) identify any significant pathway. We also specifically examined proteins coded by the genes that are adjacent to cg05917797 on the genome but found no significant associations (For log_2_ transformed protein levels, HIF1A: Beta = 0.007, SE = 0.018, t=0.398, p=0.69; PRKCH: Beta = -0.49, SE=0.026, t=-1.88, p=0.060). Examination of PRKCH in the Lothian Birth Cohort 1936 was not significant (p=0.5).

### Meta-analyses of all longitudinal dementia cohorts

To produce a more powerful analysis of epigenetic predictors of dementia risk, we meta-analyzed WHIMS and all replication cohorts (N=16,915, including 1,190 incident dementia events). cg05917797 remained significant and was the leading association (p=1.20x10^-9^). Three additional CpG sites reached epigenome-wide significance (Table 2; Supplementary Figure 5; Supplementary Table 2). These additional associations were largely driven by the other cohorts in the meta-analysis, with minimal evidence coming directly from WHIMS (all p > 0.05). For two of these CpGs, the majority of evidence came from Generation Scotland (cg17998013, chr3: 15,643,079 BP; p = 6.4x10^-9^; cg08576390, chr16:20,878,583; BP p=1.3x10^-7^). The other significant CpG was a composite of evidence from the non-WHIMS studies (cg17636193, chr11:101,785,595BP, p = 4.6x10^-8^).

**Table 2.**
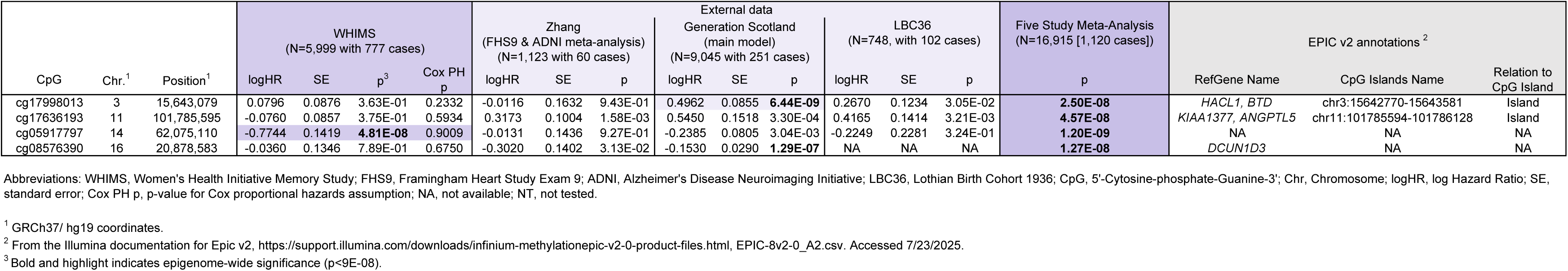
Epigenome-wide significant probes across analyses.

Pathway analyses of the top 1000 CpG sites indicated one significant KEGG pathway, “Longevity regulating pathway” (pathway containing N=88 CpGs, of which 15 were in the leading 1000 CpG sites of the data, p=1.08x10^-4^, FDR=0.04).

## DISCUSSION

In this EWAS among 5,999 older women, we identified cg05917797 as a prospective, blood-based epigenetic marker of dementia developed up to 25 years later where higher methylation was associated with lower dementia risk. The association was supported by stability across sensitivity analyses, replication in independent cohorts, and directionally concordant associations with Braak pathology in brain-based EWAS. The association also persisted after adjustment for *APOE* e4 allele status, plasma p-tau217, and epigenetic clocks, suggesting that it may capture a risk axis not fully explained by these established markers.

External epigenetic studies of brain tissue provide additional support and potential biological context for cg05917797. The EWAS meta-analyses performed by Smith et al. identified this CpG as being associated with Braak stage in the superior/middle temporal gyrus and cerebellum, where higher methylation corresponded to lower neuropathological burden.^6^ Given that Braak staging is part of the National Institute on Aging criteria for dementia diagnosis, these findings are directionally consistent with our results. Notably, temporal lobe neurofibrillary pathology is closely linked to memory impairment in aging and Alzheimer’s disease,^21,22^ whereas the cerebellum is now recognized as participating in distributed cognitive networks rather than purely motor function.^23,24^ The absence of significant signals in the prefrontal cortex and entorhinal cortex, while potentially interesting, may simply reflect the influence of cell-type heterogeneity, sample size, and pathology across post-mortem datasets on discovery power. Notably, cg05917797 is highly methylated in blood but more intermediately methylated in brain, suggesting that reduced blood methylation in dementia cases may reflect a peripheral disease-associated regulatory state rather than a simple proxy for steady-state brain methylation.

We also examined whether cg05917797 was associated with cognitive performance, given its association with future dementia risk. Higher methylation was nominally associated with better delayed verbal recall and forward digit span in WHIMS, with directionally consistent but modest associations observed in Generation Scotland ^25^. However, cognitive testing was available only in a WHIMS subset, and adjustment for educational attainment in the primary dementia model did not materially alter the cg05917797 association. These findings suggest phenotypic overlap with cognitive measures, but do not indicate that baseline cognitive performance or educational differences explain the methylation–dementia association.

It is not clear what biology underlies the association between cg05917797 and dementia. We examined genomic annotations surrounding the locus in order to provide clues into its biological context. As indicated by positional mapping, cg05917797 lies within a regulatory region annotated as an enhancer element (GeneHancer GH14J061606) with multiple clustered interactions with adjacent genes. The CpG site overlaps long-noncoding RNA (lncRNA) LINC03033, which was not previously linked to dementia. The closest protein-coding genes are *PRKCH* (∼287KB downstream from transcription start site), recently linked to Alzheimer’s disease by GWAS,^26^ and *HIF1A* (∼87KB upstream of transcription start site), a transcription factor central to cellular responses to hypoxia that has been implicated in Alzheimer’s disease biology.^27^

We examined (and performed) mQTL GWAS to identify potential genetic drivers of methylation at this CpG and obtain positional clues for potentially relevant genes. However, the absence of significant SNP associations and negligible SNP-based heritability of cg05917797 suggest limited genetic regulation, motivating complementary evaluation of dynamic measures of gene involvement such as transcriptomic and proteomic data. Accordingly, we leveraged the concurrently measured protein data in WHIMS to achieve this but did not identify significant protein associations. Similarly, we did not identify supporting evidence from external proteomic or transcriptomic resources or transcriptomic studies.^28^

Given the overlap of cg05917797 with enhancer-associated regulatory annotations, methylation at this locus may reflect broader chromatin regulatory activity relevant to neurodegeneration. Histone acetylome studies in Alzheimer’s disease have identified widespread alterations in enhancer-associated marks such as H3K27ac across regulatory regions in brain tissue.^29,30^ Because enhancer-associated DNA methylation can mark regulatory states that also involve histone modifications, cg05917797 may index a broader epigenetic regulatory state associated with dementia risk, although further integrative analyses are required to determine whether methylation at this locus directly contributes to regulatory dysfunction.

While our EWAS identified only one epigenome-wide significant CpG, pathway analysis provides minor evidence of a broader influence of epigenetics on dementia incidence: we identified integrin signaling as significantly overrepresented among the most significantly associated CpGs. Integrin signaling links extracellular matrix organization with blood–brain barrier function and memory-related synaptic biology.^31^ In addition, pathway enrichment of the meta-analysis additionally identified the KEGG longevity regulating pathway. Given that aging is the strongest risk factor for dementia, enrichment of longevity-associated pathways may reflect epigenetic alterations occurring within broader biological systems underlying vulnerability to neurodegeneration. Although broad, this pathway captures aging-related stress-response and metabolic regulatory systems relevant to dementia.

The meta-analysis also identified additional CpG sites that showed minimal evidence of association within WHIMS alone. This may reflect heterogeneity across cohorts, including differences in sex composition, study design, follow-up duration, dementia ascertainment, methylation array content, or sampling variability. These CpGs should therefore be considered candidates for future follow-up and multi-omic characterization rather than established dementia-associated methylation markers.

Strengths of our study include the large sample size of older women with up to 25 years of follow-up for incident dementia, which was adjudicated using rigorous procedures. We replicated our analyses across four independent longitudinal cohorts, with notable diversity in cohort composition and persistence across methylation array. We leveraged multi-omic data, including brain, cognitive function, genomics, and proteomics, to provide comprehensive follow-up analysis of our main finding. More, we have meta-analyzed all cohorts, thus generating one of the most comprehensive longitudinal examinations of epigenetic predictors of dementia risk to date.

Some limitations should also be considered when interpreting these findings. First, although the association replicated across cohorts, the number of participants who developed dementia remains modest relative to typical GWAS sample sizes. This raises the possibility that additional relevant CpG associations remain undetected, beyond the single significant one that we have identified. Indeed, we may expect that methylation associations preceding disease development by decades would likely be subtle, especially if they are measured in blood, where signal may be diluted relative to brain tissue. Second, we lacked paired blood and brain methylation measurements within WHIMS, limiting our ability to determine whether disease-associated blood methylation differences at cg05917797 correspond to methylation differences in brain tissue. This also makes it more challenging to assess whether risk is symmetric around methylation levels. As blood methylation levels are very high at this CpG, it is not clear if higher levels of methylation relate to reduced risk. It may rather be that abnormally low levels of methylation relate to increased risk. Third, although our longitudinal evaluation suggests that methylation at cg05917797 was associated with incident dementia, it remains possible that the signal reflects early but undetected neurodegenerative processes. Fourth, although we had a diverse sample, the majority of participants were of European ancestry, such that findings may not generalize across ancestry. Lastly, our main outcome was all-cause dementia. While this was rigorously adjudicated, it does not rule out that the observed association may be particularly relevant to a only a specific subtype of dementia; however, WHIMS did not adjudicate dementia subtypes. We note how our exploration of results in the brain focused on Braak staging, which is particularly relevant to AD-related dementia, and whose relevance to other types (e.g., trauma, infection, epilepsy, Parkinson’s disease, cerebro-vascular disease and CVD, air pollution, etc.) is unknown.

In summary, our study identified cg05917797 as a reproducible blood-based epigenetic prognostic marker of incident dementia occurring up to 25 years later. Its association with incident dementia, persistence after adjustment for established biomarkers, and concordance with brain Braak-stage EWAS support its relevance to dementia biology, while the absence of clear genetic or proteomic correlates indicates that its mechanistic basis remains unresolved.

## METHODS

### Study Population

WHIMS was an ancillary study of the WHI Hormone Therapy Trials designed to examine the effects of conjugated equine estrogen (CEE) plus medroxyprogesterone acetate vs placebo among women with an intact uterus or CEE alone vs placebo among women with hysterectomy on cognitive outcomes among 7,479 postmenopausal women.^16,17,32^ Women aged 65-79 years were recruited from 39 U.S. clinical centers during 1996-1999 and were cognitively unimpaired at enrollment. The trials were stopped in 2002 and 2004, respectively, but women continued to be followed for cognitive outcomes. Women completed annual in person cognitive assessments through 2007. In 2008, WHIMS transitioned to annual telephone-based cognitive assessments in the WHIMS-Epidemiology of Cognitive Health Outcomes (WHIMS-ECHO) study, with follow-up for cognitive outcomes through 2021.^33^ At baseline, participants completed questionnaires assessing demographic characteristics and medical history.

Among 7,479 WHIMS participants, we excluded 240 women with only one WHIMS cognitive assessment, 519 who were ineligible for dbGaP, and 304 without available baseline DNA or buffycoat, leading to 6,416 WHIMS participants whose DNA samples underwent DNAm measurement. After excluding participants who failed QC, failed fingerprinting, or were related to another participant (see below), 5,999 women remained in the analytic sample. This study was approved by the Institutional Review Board of University of California San Diego. All participants provided written informed consent.

### Methylation data

Genomic DNA samples were shipped to the University of Minnesota Genomics Center for DNAm quantification using the Infinium MethylationEPIC v2.0 kit (Illumina, Inc., San Diego, CA). Samples were assessed for quality using 17 control metrics from Illumina. Samples were removed if they did not meet Illumina’s recommended thresholds for each control metric (N=14 removed). Sex was determined by clustering samples on the average intensity values of CpG sites on the X and Y chromosomes.^34^ Samples that fell outside of the female cluster were excluded (N=8 removed). Samples with mean bisulfite intensity values < 4,000 were excluded (N=1 removed). Detection p-values were calculated using out-of-band (OOB) probes.^35^ CpG measurements were set to missing if detection p > 0.05 or <= 3 detection beads (M=66,532 probes). CpG sites were removed if >5% of samples were missing data. Based on the remaining CpG sites, samples were excluded if >5% of CpG sites had missing methylation values (N=43 removed). In the remaining samples, OOB background correction, RELIC dye bias correction, and RCP probe type bias correction were applied using Enmix.^36^

### Ancestry

To determine global ancestry of each particpant, genotype data was put through a standardized SNPweights pipeline including up to 10,000 ancestry informative markers genotyped in a reference panel of 2,911 subjects from six continental groups.^37,38^ Participants were placed into three homogenous groupings using previously established cutoffs. European and European Americans (EUA; participants with ≥90% European ancestry), African and African-Americans (AFA; participants with ≥5% African ancestry, <90% European ancestry, <5% East Asian, Native American, Oceanian, and Central-South Asian ancestry; and participants with ≥50% African ancestry, <5% Native American, Oceanian, and < 10% Asian ancestry), and Indigenous American ancestry participants (IAA; participants with ≥5% Native American ancestry, <90% European, < 7% African, and <5% East Asian, Oceanian, and Central-South Asian ancestry; and participants with ≥60% Native American ancestry, <20% East Asian, <15% Central-South Asian, and <5% African and Oceanian ancestry).

For N=893 samples, genotype data were not available; therefore, we derived ancestry from methylation data. We used the nmode function in Enmix to identify 2,072 CpG sites with a trimodal distribution, under the assumption that these CpG sites would be highly enriched for measuring SNPs. PCs were estimated using these CpG sites. To identify which PCs were associated with ancestry, we measured the correlation between each PC and the PC values projected from the SNP-based global ancestry panel. Two methylation-SNP derived PCs were significantly correlated (PC2 to ancestry PC1 r = 0.81; PC4 to ancestry PC2 r= 0.43) to the first two PCs from the global ancestry panel. Using the samples with known, classified genetic ancestry, ellipsoids were drawn around population means. Unclassified samples were then classified as European, African, or Latin American ancestry, depending upon which ellipsoid they fell into.

### Principal components

A random subsample of 172,000 CpG sites spanning the genome was extracted. Methylation values were transformed to m-values. Missing CpG data were imputed to the mean. Principal components were calculated in unrelated, non-replicate samples using the big_randomSVD function in the bigstatsr library.^39^ In remaining samples, PC values were estimated via projection.

### Sample Concordance

Concordance between samples was measured using the SNP fingerprinting probes built into the array. Illumina SNP probes were converted to genotype data. These genotypes were assessed for pairwise identity-by-descent (IBD) in PLINK. If a sample had a 100% match with a non-replicate sample, the pair was compared against genotype array data. Under the assumption that array genotypes represented the ‘truth’ dataset, the sample from the pair that did not match the array genotype was excluded.

### Outcome phenotype assessment

Our primary outcome was incident all-cause dementia. Details on dementia adjudication have been published.^16,17^ Briefly, participants completed the Modified Mini-Mental State Examination, with those scoring below specific cut points (based on age and education) completing a modified Consortium to Establish a Registry for AD battery of neuropsychological tests and standardized questionnaires in person. A local site physician with expertise in dementia diagnosis classified women as having no dementia or probable dementia based on *Diagnostic and Statistical Manual of Mental Disorders, Fourth Edition (DSM-IV)* criteria. All data were sent to the WHIMS Clinical Coordinating Center for review and central adjudication of final diagnosis by a panel consisting of a neurologist, geriatric psychiatrist, and geropsychologist. WHIMS-ECHO used a validated protocol of telephone-based cognitive assessments and informant interviews, and a similar protocol to that of WHIMS for ascertainment and central adjudication of final diagnosis.^33^

### EWAS

In WHIMS, the association between DNA methylation and time to incident dementia was assessed using Cox proportional hazards regression models. Women were followed from the date of WHIMS randomization. Time was defined as the date of the cognitive assessment that triggered the first diagnosis of dementia, loss to follow-up, or November 3, 2021 (end of study), whichever came first. Women who did not develop MCI or dementia, including those who died or ended the study for other reasons, were censored at the date of their final cognitive assessment, consistent with prior WHIMS publications.^16,17^ Our model included covariates for CpG site (as m-values), age at baseline, cell type proportions (B, CD4T, CD8T, NK, monocytes, neutrophils), methylation derived PCs 1-10, methylation ancestry PCs 1-2, and chip-row position (as a factor). We used Schoenfeld residuals to evaluate proportional hazards assumptions. Statistical significance was declared at p = 9 x10^-8^ and suggestive association was declared arbitrarily at 5x10^-6^.^40^

As an initial sensitivity analysis, the EWAS was repeated in the subset of participants of European ancestry. As a second sensitivity analysis, we adjusted for potential confounders, including HT arm, educational attainment, and smoking history, in addition to the variables included in the base model. For our leading CpG site, we also evaluated the association where the Cox model was additionally adjusted for dementia biomarkers, including *APOE* e4 allele status, epigenetic clock based measures of age acceleration (PCGrimAge and AgeAccelGrim2; results previously detailed),^41^ and plasma p-tau217 (log_2_ transformed), in the subsets of participants for whom these variables had been assessed. Plasma p-tau217 was measured at baseline using the ALZpath Simoa pTau-217 v2 assay, as previously described.^42^ Finally, we accounted for the competing risk of death of the leading CpG site using a cause-specific competing risks Cox model. Gene set enrichment analysis was performed using missMethyl (*gometh*) (PMID: 26424855)^43^, applying our top 1,000 CpG list as the foreground and all tested CpGs as the background, with bias correction (prior.prob = TRUE) to account for probe-per-gene differences and multi-gene CpG annotations.

### Cognitive Assessments

A subset of WHIMS participants was part of the Women’s Health Initiative Study of Cognitive Aging (WHISCA). WHISCA participants were enrolled on average three years after WHIMS enrollment and completed an additional comprehensive cognitive assessment annually through 2010. Test battery and protocol details are published elsewhere.^44^ The cognitive battery included tests of global cognition (3MS score [Modified Mini-Mental State]), verbal knowledge (Primary Mental Abilities Vocabulary total correct – 1/3 incorrect), verbal fluency (letter fluency), figural memory (Benton Visual Retention Test: total figures with errors), verbal memory (California Verbal Learning Test [CVLT], CVLT-Long Delay, CVLT Short Delay), attention and working memory (Digit Span forward, Digit Span backward), spatial ability (Card Rotation Test [CRT]: total correct – total incorrect), and fine motor speed (Finger Tapping Test Total dominant + non-dominant hand).

Associations between methylation and a battery of cognitive variables were evaluated using linear mixed models, adjusting for days to battery, form version, age at baseline, cell type proportions, PCs 1-10, ancestry PCs, and chip-row position, and included a subject specific intercept random effect term to account for the repeated measures. To evaluate whether associations were due to educational attainment, a second model was evaluated that included educational attainment as an additional covariate. Time interaction models were evaluated using the aforementioned model, with an additional term added for the interaction of methylation and days to battery.

### PWAS

Plasma specimens (55ml) obtained from the WHIMS cohort were analyzed by SomaScan proteomics analysis using 96-well plates on a Tecan Fluent 780 liquid handling robot at the BIDMC Genomics, Proteomics, Bioinformatics and Systems Biology Center using the 11k SomaScan v5.0 Assay Kit for human plasma that measures expression of 10,778 proteins using highly selective single-stranded modified Slow Off-rate Modified DNA Aptamers (SOMAmer) according to the manufacturer’s standard protocol (SomaLogic; Boulder, CO). Five pooled plasma Calibrator replicates, 3 pooled plasma Quality Control replicates, and 3 buffer replicates were used to control for batch effects and to estimate accuracy, precision, and buffer background over time. Twelve hybridization controls and 296 non-human SOMAmers were added alongside the 10,778 SOMAmers to control for readout variability for each set of 85 test samples. Sample to sample variability was further controlled by several hybridization spike-in controls. Hybridization normalization, plate scaling, median normalization, calibration, and Adaptive Normalization by Maximum Likelihood (ANML) of the SomaScan data, performed according to the standard quality control (QC) protocols at SomaLogic, demonstrated that all samples passed the established QC criteria and were fit for further analysis. Protein values were log2 transformed. The association between cg05917797 and protein abundance was measured using linear regression with cg05917797 as the outcome variable, including covariates for age at baseline, cell type proportions, PCs 1-10, ancestry PCs, and chip-row position.

### Replication cohorts

Details on the replication cohorts, including study population, DNAm measurement and quality control, and dementia ascertainment, are described in the Supplementary Methods.

#### Generation Scotland (GS)

GS consisted of 9,045 individuals; 2.8% (251 subjects) developed dementia during the study period. Participants were excluded if they were 65 years or younger at the time of dementia or censoring. EWAS conducted on m-values were adjusted for age, estimated cell proportions (B, CD4+ T, CD8+ T, eosinophils, NK, monocytes), DNAm PCs 1-20, sex, and batch. Mixed effect Cox models are required to adjust for relatedness in GS. However, due to the computation intensity of these models, Cox proportional hazards were first run to identify borderline significant CpGs (p<1x10^-4^, n=327). These CpGs were then used in mixed effects Cox models, which adjust for the effects of ID and batch.

#### Lothian Birth Cohort 1936 (LBC1936)

LBC1936 consisted of 748 individuals; 13.6% (102 subjects) developed dementia during the study period. EWAS conducted on m-values were adjusted for age, measured cell counts (basophils, eosinophils, lymphocytes, monocytes, neutrophils), DNAm PCs 1-20, sex, and batch.

#### Alzheimer’s Disease Neuroimaging Initiative (ADNI) and Framingham Heart Study Exam 9 (FHS9) cohorts

Zhang et al. recently published a meta-analysis of incident dementia in ADNI and FHS9.^15^ ADNI consisted of 216 individuals; 8.3% (18) developed dementia. FHS9 consisted of 907 individuals; 4.6% (42) developed dementia. The ADNI EWAS was conducted on beta-values, and adjusted for age, PCs 1-2 of immune cell-type proportions, and sex. The FHS9 EWAS was conducted on beta-values and adjusted for age, immune cell-type proportions (B, CD4T, granulocytes, monocytes, NK), and sex.

### EWAS Meta-analysis

We performed a meta-analysis of EWAS for incident all-cause dementia among five independent prospective cohorts (N=16,915 men and women): WHIMS, FHS9, ADNI, GS, and LBC1936. EWAS were fixed effects sample-size weighted meta-analyzed using the square root number of incident dementia events as weights. Only CpGs present in one or more datasets were included.

### GWAS of cg05917797

WHIMS participants were genotyped using Illumina genotyping arrays. Samples and variants underwent standard quality control and were imputed to the Haplotype Reference Consortia panel using the RICOPILI pipeline.^45^ The association between cg05917797 methylation and genotype at each SNP was assessed using additive linear regression models, adjusted for ten genomic PCs, in PLINK2.^46^ Only common SNPs (MAF > 1%) were considered. SNP based heritability was calculated from GWAS summary data using LD score regression^47^ and 1000Genomes Phase 3 European LD reference data.

## Data Availability

All data produced in the present study are available upon reasonable request to the authors

https://www.whi.org/doc/PP-policy.pdf

## ACKNOWLEDGEMENTS

We thank the WHI participants, staff, and investigators. The short list of WHI investigators can be found at: https://www-whi-org.s3.us-west-2.amazonaws.com/wp-content/uploads/WHI-Investigator-Short-List.pdf. The full list of WHI Investigators can be found at the following site: https://www.whi.org/doc/WHI-Investigator-Long-List.pdf

## AUTHOR CONTRIBUTIONS

A.X.M. and A.H.S. contributed to the study concept and design. A.H.S. obtained funding. A.X.M. wrote the first draft of the manuscript. A.X.M. and C.M. conducted the statistical analyses. T.L. led the collection of SomaScan data. A.X.M. and C.M. had full access to all the WHIMS data in the study and take responsibility for the integrity of the data and the accuracy of the data analysis. A.X.M. and C.M. accessed and verified the WHIMS data. J.A.R. and R.E.M. led replication analyses in the Generation Scotland and Lothian Birth cohorts. S.E.H. and M.P. led replication of SomaScan findings. All authors provided input to the interpretation of results, revised and approved the final manuscript, and had final responsibility for the decision to submit for publication.

## DATA SHARING STATEMENT

De-identified data from the study and supporting documents can be made available after publication to researchers with investigator support, after approval of a proposal by the WHI Publications and Presentations Committee and with a signed data access agreement (see https://www.whi.org/doc/PP-policy.pdf). Please contact Aladdin Shadyab

