## supplementary methods and figures for "Blood DNA Methylation Predicts Long-Term Risk of Dementia in Prospective Cohorts"

**Generation Scotland**

Generation Scotland comprises approximately 24,000 volunteers with socio-demographic, clinical, genetic, methylation and proteomic data and linkage to medical records.^1^ Participants were recruited from across Scotland between 2006 and 2011 when they were aged between 17 and 99 years. Blood samples were taken during the initial clinic visit (N~20,000), together with lifestyle, cognitive and health questionnaires. Informed consent for electronic-health records linkage for both primary and secondary care data was provided. For the 18,869 individuals with methylation data, the mean age is 47.12 and 58.8% are female.^2^

**Ethics**

Ethical approval was received from the NHS Tayside Committee on Medical Research Ethics (REC Reference Number: 05/S1401/89) and Research Tissue Bank status was granted by the East of Scotland Research Ethics Service (REC Reference Number: 20/ES/0021). Participants provided written informed consent.

**DNA methylation**

Baseline blood samples were processed for DNA methylation measurements. DNA was treated with sodium bisulphite and methylation measured using the Illumina Infinium HumanMethylationEPIC BeadChip array v1.0.^3^ Full details of sample processing and quality control (QC) have been described previously.^2^ DNAm was profiled in four separate sets (Post QC: N_Set1_ = 5,087, N_Set2_ = 459, N_Set3_ = 4,450, N_Set4_ = 8,873, Total N = 18,869).^2^ Samples were removed if the median methylated signal intensity was more than three standard deviations lower than expected, if self-reported sex differed from methylation-derived sex, and if > 1% CpGs had a detection P > 0.05 (set 1) or if > 0.5% CpGs in the sample had a detection P > 0.01 (sets 2 – 4). Probes were removed if the >0.5% of the samples had a detection P >0.05 (set 1) or if >1% samples had a detection P > 0.01 (sets 2-4) or if the beadcount was less than 3 in >5%. 18,869 samples and 752,722 CpG sites were retained following quality control. Normalised methylation M-values were used for downstream analyses.

**Dementia Ascertainment**

In the Generation Scotland cohort, dementia diagnosis was obtained by linkage to primary and secondary care records (available for 7,580 and 21,725 individuals respectively), using CALIBER/HDRUK consensus definitions.^4^ The first record of an event was used to define the date of diagnosis. For the Cox model analysis, the censor date was set to October 2024 – the latest date for which secondary care data were available. Participants were filtered to those aged over 65 at diagnosis or censor to align with other data in the meta-analysis and to remove any early-onset dementia diagnoses. Any prevalent dementia cases were also removed, leaving 9,045 individuals with methylation data (251 cases and 8,794 controls) for downstream modelling. All analysis was undertaken in R v.4.4.2.

**Cox models**

Time to dementia diagnosis (maximum follow-up 18.6 years) was modelled for each CpG (N = 752,722), adjusting additionally for age, sex, methylation measurement batch, estimated white cell proportions, and the first 20 methylation principal components. Mixed-effect Cox models to account for relatedness as a random effect were required due to related individuals being present within Generation Scotland. However, due to the large number of CpGs to be modelled, the models were first run as Cox proportional hazard models (survival package v.3.8.3) without adjusting for relatedness to reduce the computational intensity. Subsequently, any CpGs with P < 1x10^-4^ (N = 327) were run through mixed-effects cox-models (coxme package v.2.2.22), where adjustment for relatedness was carried out using a kinship matrix (kinship2 package v. 1.9.6.1). The models are outlined below:

**Coxph:** Time-to-dementia diagnosis ~ CpG + age + sex + Batch + estimated white cell proportions + 20 methylation principal components

**Coxme:** Time-to-dementia diagnosis ~ CpG + age + sex + (1|id) + (1|Batch) + estimated white cell proportions + 20 methylation principal components

**Lothian Birth Cohort 1936**

**LBC1936**

The Lothian Birth Cohort 1936 is a longitudinal study consisting of individuals residing within the Lothian region of Scotland who were born in 1936, undertook the Moray House Test No.12 aged around 11 years while at school in Scotland in 1947, and were invited to join the LBC1936 aged ~70.^5^ Participants underwent physical, cognitive and medical assessments and provided blood samples at age 70 (wave 1) and approximately triennially thereafter.^6^ Data from wave 2 (mean age 72.5, 48% female) were used for this analysis (n = 895).

**Ethics**

Ethical approval for the LBC1936 study was obtained from the Lothian Research Ethics committee (LREC/2003/2/29) theMulti-Centre Research Ethics Committee for Scotland (MREC/01/0/56), and the Scotland A Research Ethics Committee (07/MRE00/58). All participants gave written informed consent for their data to be used for health research.

**DNA methylation**

DNA methylation was measured using DNA extracted from whole blood samples, via the Illumina 450K methylation array at the Edinburgh Clinical Research Facility. Quality control steps have been described in depth previously.^7^ Briefly, internal controls were used to background-correct and normalise raw-intensity data. Low quality samples were removed by manual inspection for inadequate hybridisation or nucleotide extension, or issues related to bisulphite conversion and staining signal. Probes with a low detection rate (<95% at P < 0.01) were removed and samples with a low call rate (<450,000 probes detected at P<0.01) were also removed. Any samples with mismatched genotype and SNP control probes or incorrect DNA methylation-predicted sex were also removed.

**Dementia Ascertainment**

Consent for health-record linkage is available for participants in wave 2 onwards. Dementia diagnosis was ascertained through a clinical consensus meeting following electronic health record (EHR) review and, where any signs of mild cognitive impairment were present or possibility of dementia raised by the participant themselves or LBC1936 researcher, a clinician home visit was undertaken for further assessment (https://pmc.ncbi.nlm.nih.gov/articles/PMC10152609/).

The consensus meeting classed each potential case as dementia, probable dementia, possible dementia or no dementia. Diagnosis dates were only available for those with a definitive dementia diagnosis and therefore those with mild cognitive impairment (N = 13), possible dementia (N = 7) or without consent for linkage (N = 1) were removed from subsequent analyses. Six additional individuals were removed from analysis where a retrospective diagnosis was made pre-baseline (N = 2), there were inconsistent dates (N = 1) or linkage was unavailable (N = 3). 765 individuals remained at this stage (104 cases, 661 controls).

**Cox models**

For the purposes of Cox models, an additional 17 individuals were removed from analysis due to missing data for measured white cell counts (N = 12) and where methylation set inclusion prevented model convergence (N = 5). 748 individuals were taken forward for Cox modelling (102 cases, 646 controls). Time to dementia diagnosis was modelled for each CpG (N = 459,309) with adjustment for age, sex, methylation measurement set, white cells as measured by clinical haematology (neutrophils, lymphocytes, monocytes, eosinophils, basophils) and the first 20 methylation principal components.

**Framingham Heart Study**

The Framingham Heart Study (FHS) is a community-based study examining risk factors for cardiovascular disease in Framingham, Massachusetts.^8^ The FHS Offspring cohort included participants from the second generation and their spouses. Blood samples were collected from the FHS Offspring cohort during 2011-2014 at Exam 9. All 907 participants in FHS examined in this study were non-Hispanic White and free of dementia at Exam 9.

**DNA methylation**

DNAm was measured using the Illumina HumanMethylationEPIC v1 bead chip. Quality control steps for both cohorts were described previously.^9^ Briefly, probes with a detection p-value <0.01 in ≥90% of the probes were selected. Probes that start with “cg,” were next selected. Probes that met the following criteria were removed using the function fmSNPandCH from the DMRcate R package: located on X and Y chromosomes, are cross-reactive, or located close to single nucleotide polymorphism.

Samples with bisulfite conversion rate <85% were removed, as were those for which the DNAm predicted sex status differed from the recorded sex status. Sex prediction was determined using the getSex function from the minfi R package. Outliers were identified using principal component analysis (PCA) among the 50,000 most variable CpGs. Samples that were outside the range of ±3 standard deviations from the mean of PC1 and PC2 were excluded.

Data were subsequently normalized using the dasen method using the wateRmelon R package.^10^ The EpiDISH R package was used to estimate immune cell type proportions (B lymphocytes, natural killer cells, CD4+ T cells, CD8+ T cells, monocytes, neutrophils, and eosinophils). Batch effects from methylation plates were corrected using the BEclear R package.^11^

**Dementia Ascertainment**

In FHS, participants underwent a Mini-Mental State examination at each exam. Participants completed a 45-minute neuropsychological test every 5 to 6 years since 1999.^9^ Participants who were flagged for possible cognitive impairment according to these assessments were subsequently invited to complete further annual neurological and neuropsychological evaluations. Participants who showed improvement at two consecutive annual evaluations returned to the regular follow-up schedule. Dementia ascertainment is described elsewhere.^12^ Dementia was ascertained by a review panel based on the Diagnostic and Statistical Manual of Mental Disorders, fourth edition (DSM-IV) criteria.

**Cox models**

Cox proportional hazards regression analyses were performed using the coxph function in the survival R package, as previously described.^9^ For the FHS dataset, the following model was used: Surv (follow-up time, status) ~ methylation.beta + age + sex + immune cell-type proportions (B, NK, CD4T, Mono, Gran), where status indicates incident dementia (1=event occurred; 0=censored).

**Alzheimer’s Disease Neuroimaging Initiative**

The Alzheimer’s Disease Neuroimaging Initiative (ADNI) is a multisite, longitudinal observational study to identify validated biomarkers of AD.^13^ The earliest visit with DNAm data available was selected for each participant. All participants were cognitively unimpaired at the time of the DNAm measurement, leading to an analytic sample of 216 self-reported non-Hispanic White participants.

**DNA methylation**

DNAm was measured using the Illumina HumanMethylationEPIC v1 bead chip. Quality control steps for both cohorts were described previously.^9^ Briefly, probes with a detection p-value <0.01 in ≥90% of the probes were selected. Probes that start with “cg,” were next selected. Probes that met the following criteria were removed using the function fmSNPandCH from the DMRcate R package: located on X and Y chromosomes, are cross-reactive, or located close to single nucleotide polymorphism.

Samples with bisulfite conversion rate <85% were removed, as were those for which the DNAm predicted sex status differed from the recorded sex status. Sex prediction was determined using the getSex function from the minfi R package. Outliers were identified using principal component analysis (PCA) among the 50,000 most variable CpGs. Samples that were outside the range of ±3 standard deviations from the mean of PC1 and PC2 were excluded.

Data were subsequently normalized using the dasen method using the wateRmelon R package.^10^ The EpiDISH R package was used to estimate immune cell type proportions (B lymphocytes, natural killer cells, CD4+ T cells, CD8+ T cells, monocytes, neutrophils, and eosinophils). Batch effects from methylation plates were corrected using the BEclear R package.^11^

**Dementia Ascertainment**

Dementia ascertainment in ADNI is described elsewhere.^14^ Subjects with AD had an MMSE score between 20 and 26 (inclusive), with a Clinical Dementia Rating score of 0.5 or 1. For memory criterion, delayed recall of 1 paragraph from the Logical Memory II subscale of the Wechsler Memory Scale-Revised was used with cutoff scores based on education. Participants with AD had scores ≤8 for 16 years of education, ≤4 for 8-15 years of education, and ≤2 for 0-7 years of education. Participants with AD had to meet the National Institute of Neurological and Communicative Disorders and Stroke-Alzheimer’s Disease and Related Disorders Association criteria for probable AD.

**Cox models**

Cox proportional hazards regression analyses were performed using the coxph function in the survival R package, as previously described.^9^ For ADNI, only the two first principal components of immune cell-type proportions, PC1 and PC2, were included, given the smaller sample size; PC1 and PC2 explained 90% of the variance in estimated immune cell-type proportions. The following model was fitted: Surv (follow-up time, status) ~ methylation.beta + age + sex + PC1 + PC2.

**Five Supplementary Figures**

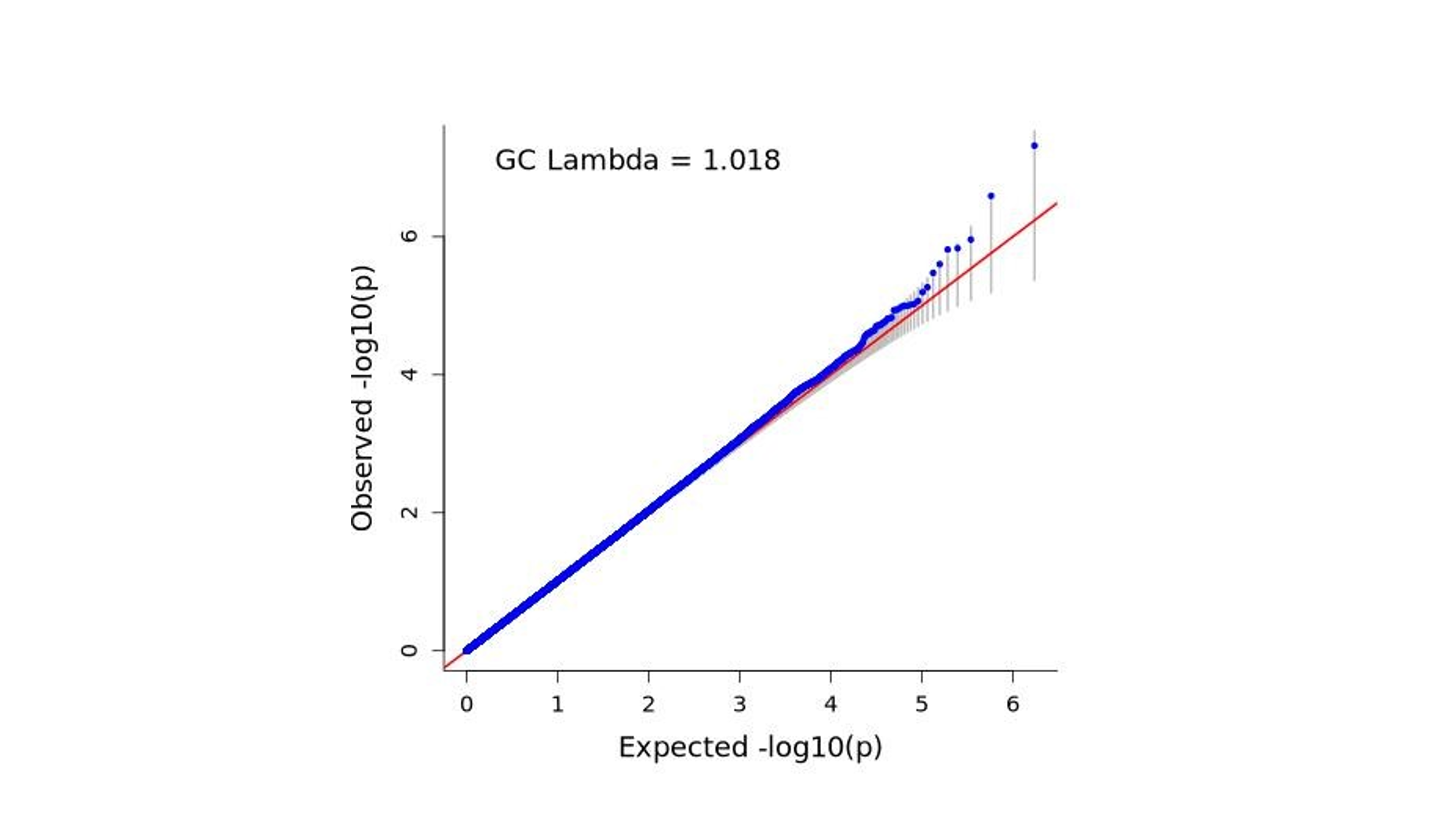

**Supplementary Figure 1.** QQ plot of dementia EWAS in WHIMS.

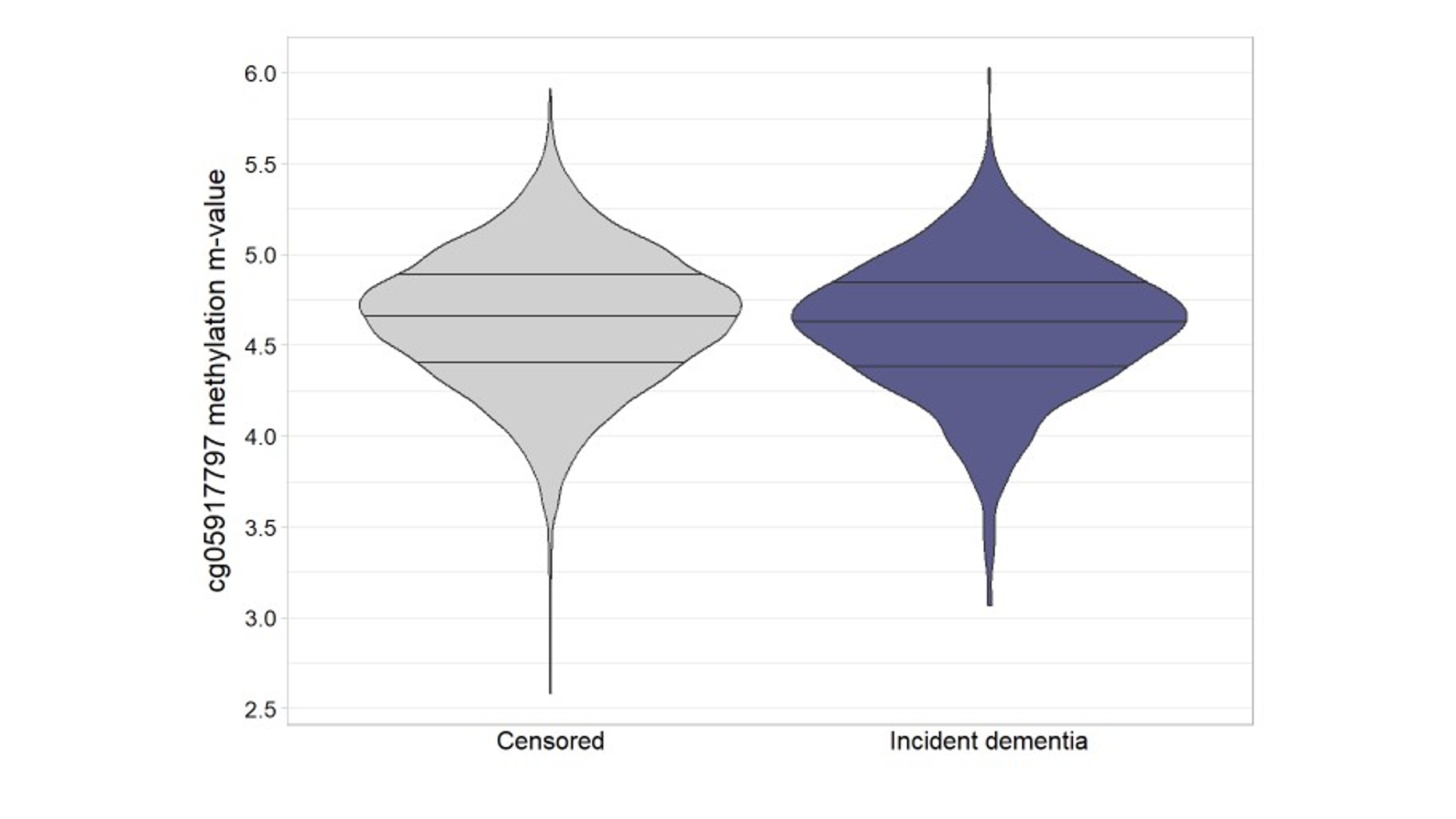

**Supplementary Figure 2.** Violin plot of cg05917797 methylation by dementia status. Horizontal bars represent the methylation m-value quartiles.

(N=5,996 WHIMS subjects)

*Note: N=5,999 for the overall WHIMS EWAS. 3 subjects were missing data for cg05917797*

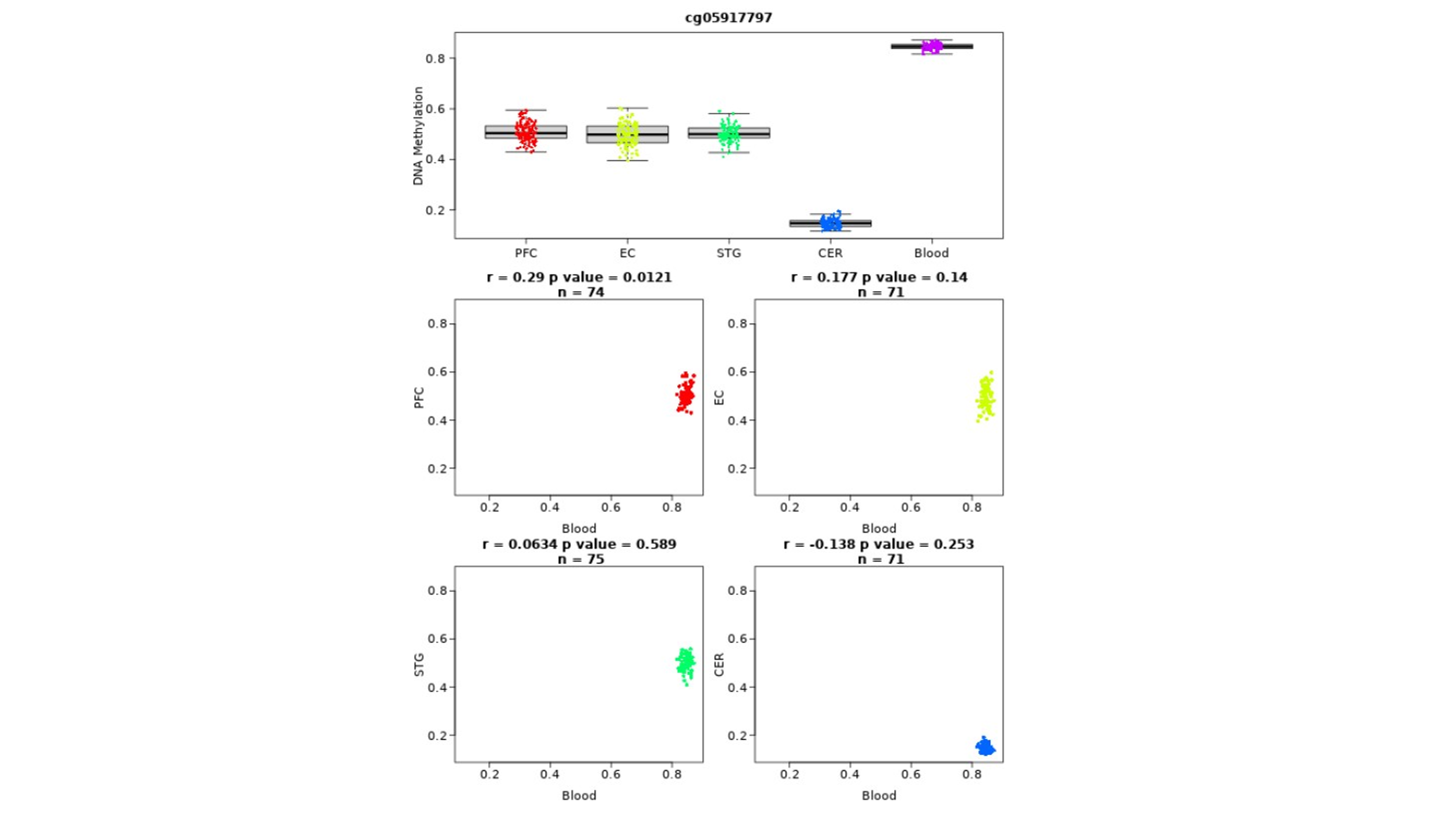

**Supplementary Figure 3.** Scatter plots of cg05917797, as measured by the Blood Brain DNA Methylation Comparison Tool. Each panel represents a different brain tissue type where correlation between blood and brain were assessed. The x-axis of each panel depicts blood measures of methylation, the y-axis depicts the corresponding brain-based measure. Abbreviations: PFC, prefrontal cortex; EC, entorhinal cortex; STG, superior temporal gyrus; CER, cerebellum.

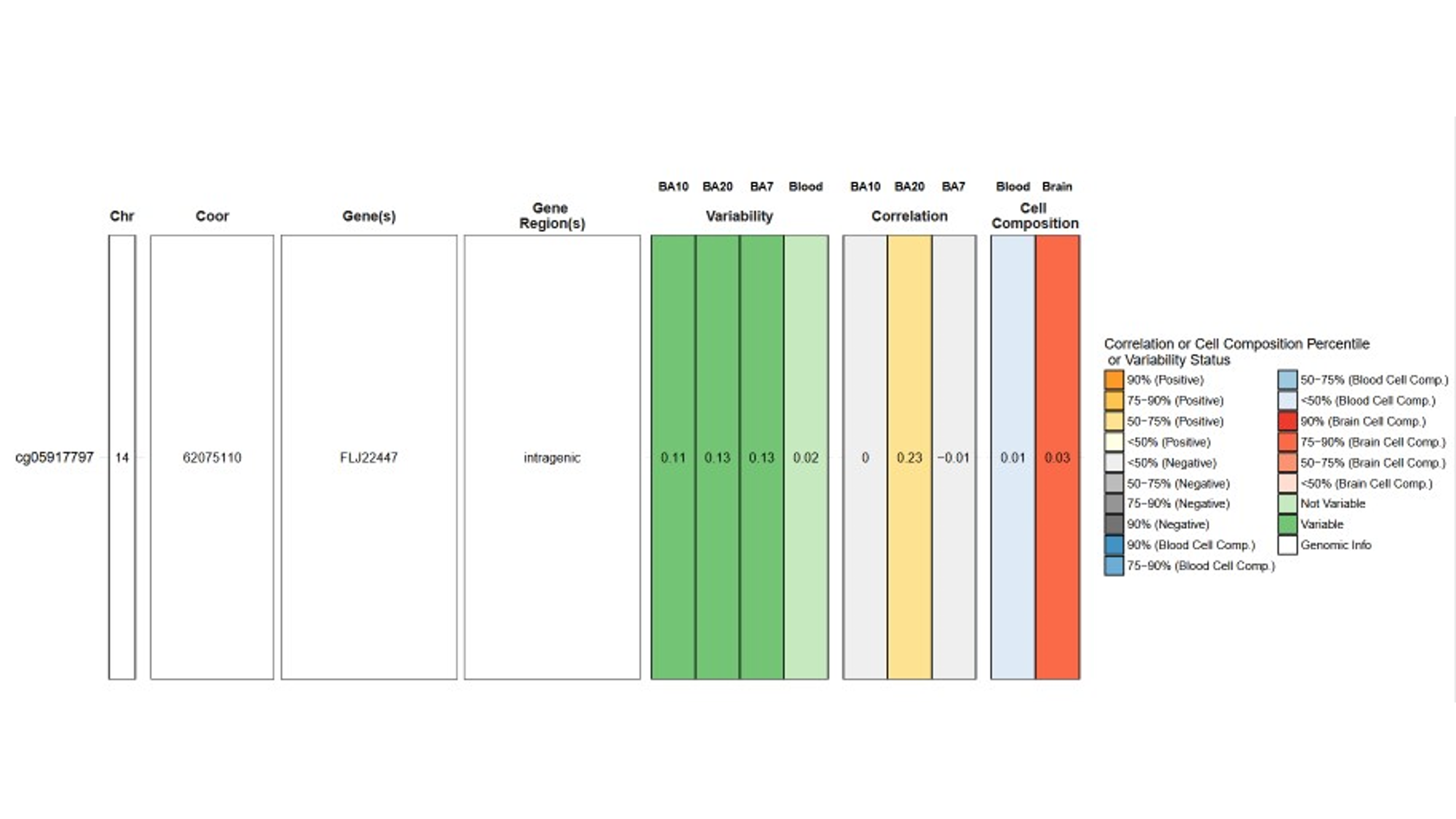

**Supplementary Figure 4**. BECON tool results for cg05917797.

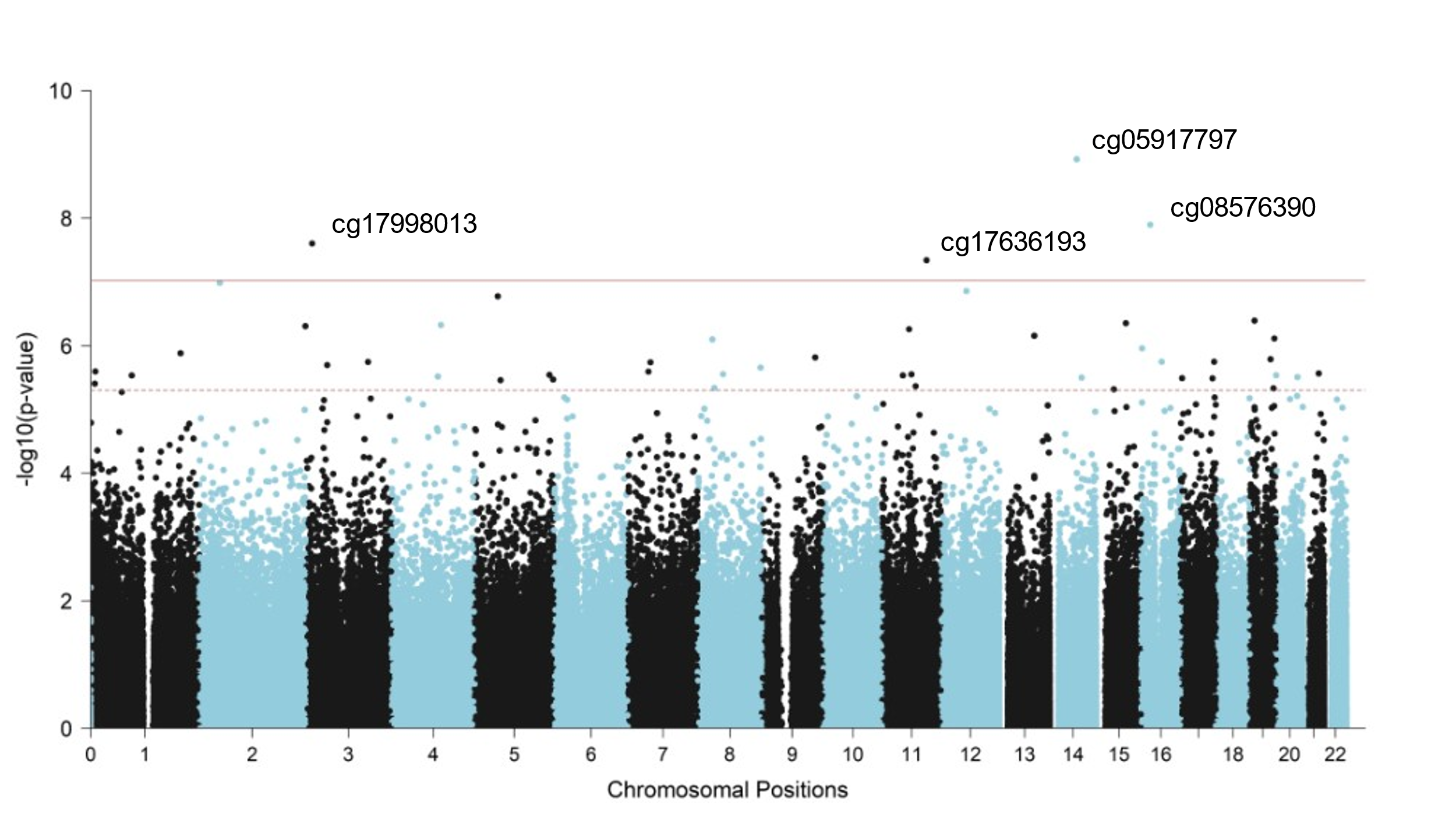

**Supplementary Figure 5.** Manhattan plot of incident dementia EWAS meta-analysis. The solid red line indicates genome-wide significance (p < 9x10^-8^) and the dotted red line indicates suggestive association (p < 5x10^-6^)
